# Clinical, neuroimaging and molecular spectrum of *TECPR2*-associated hereditary sensory and autonomic neuropathy with intellectual disability

**DOI:** 10.1101/2020.10.10.20202622

**Authors:** Sonja Neuser, Barbara Brechmann, Gali Heimer, Ines Brösse, Susanna Schubert, Lauren O’Grady, Michael Zech, Siddharth Srivastava, David A. Sweetser, Yasemin Dincer, Volker Mall, Juliane Winkelmann, Christian Behrends, Basil T Darras, Robert J Graham, Parul Jayakar, Barry Byrne, Bat El Bar-Aluma, Yael Haberman, Amir Szeinberg, Hesham Mohamed Aldhalaan, Mais Hashem, Amal Al Tenaiji, Omar Ismayl, Asma E. Al Nuaimi, Karima Maher, Shahnaz Ibrahim, Fatima Khan, Henry Houlden, Vijayalakshmi Salem Ramakumaran, Alistair T Pagnamenta, Jennifer E Posey, James R Lupski, Wen-Hann Tan, Gehad ElGhazali, Isabella Herman, Tatiana Muñoz, Gabriela M. Repetto, Angelika Seitz, Mandy Krumbiegel, M. Cecilia Poli, Usha Kini, Stephanie Efthymiou, Jens Meiler, Reza Maroofian, Fowzan S. Alkuraya, Rami Abou Jamra, Bernt Popp, Bruria Ben-Zeev, Darius Ebrahimi-Fakhari

**Author notes:** <u>Correspondence to:</u> Corresponding author: Sonja Neuser, MD, Corresponding author’s address: Philipp-Rosenthal-Straße 55, Institute of Human Genetics, University of Leipzig Medical Center, 04103 Leipzig, Germany. authors contributed equally to this work (Sonja Neuser, Barbara Brechmann, Gali Heimer, Bernt Popp, Bruria Ben-Zeev, Darius Ebrahimi-Fakhari).

## Abstract

**PURPOSE:** Bi-allelic *TECPR2* variants have been associated with a complex syndrome with features of both a neurodevelopmental and neurodegenerative disorder. Here, we provide a comprehensive clinical description and variant interpretation framework for this genetic locus.

**METHODS:** Through an international collaboration, we identified 17 individuals from 15 families with bi-allelic *TECPR2*-variants. We systemically reviewed clinical and molecular data from this cohort and 11 cases previously reported. Phenotypes were standardized using Human Phenotype Ontology terms.

**RESULTS:** A cross-sectional analysis revealed global developmental delay/intellectual disability, muscular hypotonia, ataxia, hyporeflexia, respiratory infections and central/nocturnal hypopnea as core manifestations. A review of brain MRI scans demonstrated a thin corpus callosum in 52%. We evaluated 17 distinct variants. Missense variants in *TECPR2* are predominantly located in the N- and C-terminal regions containing β-propeller repeats. Despite constituting nearly half of disease associated *TECPR2* variants, classifying missense variants as (likely) pathogenic according to ACMG criteria remains challenging. We estimate a pathogenic variant carrier frequency of 1/1,221 in the general and 1/155 in the Jewish Ashkenazi populations.

**CONCLUSION:** Based on clinical, neuroimaging and genetic data, we provide recommendations for variant reporting, clinical assessment, and surveillance/treatment of individuals with *TECPR2*-associated disorder. This sets the stage for future prospective natural history studies.

**CONFLICTS OF INTEREST:** All authors involved in the study declare no conflicts of interest relevant to this study.

## INTRODUCTION

TECPR2 belongs to the tectonin β-propeller repeat-containing protein family and is implicated in the autophagy pathway. (Oz-Levi, Gelman, Elazar, & Lancet, 2013; Stadel et al., 2015) Autophagy is critical to the development and function of the central nervous system. Loss-of-function variants in several genes of the autophagy pathway lead to both neurodevelopmental and neurodegenerative diseases. (Ebrahimi-Fakhari et al., 2016; Menzies et al., 2017; Teinert, Behne, Wimmer, & Ebrahimi-Fakhari, 2019)

In 2012, Oz-Levi et al. identified the homozygous *TECPR2* variant c.3416del, p.(Leu1139Argfs*75) in five individuals from three Jewish Bukharian families and classified the syndrome as a novel subtype of hereditary spastic paraplegia (HSP) (SPG49; MIM# 615000).(Oz-Levi et al., 2012) To date, 11 individuals with bi-allelic *TECPR2* variants have been reported. (Covone et al., 2016; Heimer et al., 2016; Oz-Levi et al., 2012; Patwari, Wolfe, Sharma, & Berry-Kravis, 2020; Zhu et al., 2015) All individuals showed muscular hypotonia and most had global developmental delay followed by intellectual disability. Only a subset of individuals displayed progressive spasticity as a characteristic HSP symptom. An autonomic and sensory neuropathy with respiratory, gastrointestinal and cardiovascular system involvement was present in a subset of individuals and central apnea was found to account for a large part of the morbidity. (Heimer et al., 2016; Patwari et al., 2020)

Beside two founder variants (c.3416del, p.(Leu1139Argfs*75) in the Jewish Bukharian background and c.1319del, p.(Leu440Argfs*19) in the Jewish Ashkenazi background), likely derived as new variants under a Clan Genomics hypothesis (Lupski, Belmont, Boerwinkle, & Gibbs, 2011), two other truncating and three missense *TECPR2* variants have been associated with the disease. Expression analyses in cell lines transfected with the p.(Leu1139Argfs*75) variant indicated escape from nonsense mediated RNA-decay (NMD) but degradation of the truncated protein. (Oz-Levi et al., 2012) Functional data is largely missing for other described variants. This poses challenges for the interpretation of missense variants, for which normal expression of an altered protein is expected. All variants have been reported based on clinical overlap but have yet to be scored through the five-tier variant classification system recommended by the American College of Medical Genetics and Genomics (ACMG). (Richards et al., 2015) The lack of functional data, and reliable variant classification have prevented an estimation of carrier frequencies and disease incidence, genotype-phenotype correlation analyses and the ability to make a genetic diagnosis in novel cases.

Through an international collaboration, we assembled a cohort of 28 individuals from 24 families of different ethnic backgrounds with known/novel disease-associated *TECPR2*-variants. Based on a detailed review of the published cases and comparison with the herein described individuals, we provide a systematic quantitative clinical synopsis based on Human Phenotype Ontology (HPO). (Köhler et al., 2019) We provide recommendations for clinical management including surveillance and symptomatic treatment. An annotation and classification of all disease-associated variants according to the current ACMG recommendations is provided. (Richards et al., 2015) Using public databases, we estimate carrier frequencies and disease incidence. Based on this curated phenotype and genotype dataset, we propose a framework for reporting and validating *TECPR2* variant alleles.

## MATERIALS AND METHODS

### Editorial Policies and Ethical Considerations

The study adheres to the principles set out in the Declaration of Helsinki. The following Research Ethics Committee approved genetic testing in research setting within the study: Ethical Committee of the Medical Faculty, Leipzig University (P1), Institutional Review Board at Boston Children’s Hospital (IRB-P00033016; P2, P4 and P5), Ethics Review Board of Technical University of Munich (P3), Institutional Review Board of King Faisal Specialist Hospital and Research Center (KFSRHC RAC# 2080006 and 2121053; P7, P8 and P13), Institutional Review Board at University College London (P14 and P15, SYNaPS cohort), East of England and South Cambridge Research Ethics Committee (REC: 14/EE/1112) for 100,00 Genomes Project Protocol (P16), Institutional Review Board at Baylor College of Medicine (H-29697) and Comité Etico Cientifico at Facultad de Medicina, Clinica Alemana Universidad del Desarrollo (P17). Genetic testing for P6, P9, P10, P11 and P12 was performed in a diagnostic setting. The authors received and archived written consent of the legal guardians to publish genetic and clinical data (P1 −P17) as well as photographs, computed tomography (CT) scan and magnetic resonance imaging (MRI) images (P1, P4, P6, P11, P13, P15, P16, P17).

### Cohort

All 17 individuals described herein (P1 – P17) were recruited through GeneMatcher (Sobreira, Schiettecatte, Valle, & Hamosh, 2015) or personal communication, from different institutions in Germany, Israel, United States, Saudi Arabia, the United Arab Emirates, Great Britain, Pakistan and Chile. Genotypic data from P3 and P13 were previously reported without a detailed clinical description (P3: reported as CB-DYS-125 in (Zech et al., 2020); P13: reported as 09DG00835 (Shams Anazi et al., 2017)).

### Clinical Spectrum

Molecular and clinical data were collected from the referring clinicians using a standardized questionnaire. All affected individuals were evaluated by a pediatric neurologist and/or geneticist. Reports of brain MRI scans were available from 15 individuals. Clinical terms were standardized using Human Phenotype Ontology (HPO) terminology. (Köhler et al., 2019) Clinical features were grouped in six categories (phenotypical abnormalities of body and face, intellectual and social development, neurological system, respiratory system, gastrointestinal system, and diagnostic procedures). Detailed case descriptions for all included individuals are provided in the File S1 and File S2 (sheet “clinical_table”).

### Genetic Analyses

Genomic DNA (deoxyribonucleic acid) was extracted using standard methods from peripheral blood samples of probands/parents. For P1, P16 and P17 conventional karyotyping was performed and all individuals, except P4, P14, P15, P16 and P17, received a chromosomal microarray. *TECPR2* variants were identified by gene panel analysis (P13), exome (P14 and P15), trio exome (P1 to P6, P10, P17), quad exome (P7 and P8), trio genome (P16) or targeted Sanger sequencing (P9, P11, P12). All herein identified *TECPR2* variants have been submitted to ClinVar (File S3 sheet “TECPR2_variants”).

### Review of Published Cases

A PubMed search identified five publications (Covone et al., 2016; Heimer et al., 2016; Oz-Levi et al., 2012; Patwari et al., 2020; Zhu et al., 2015) describing 11 individuals from nine families diagnosed with *TECPR2*-associated disease (searched on 2020-09-10). Phenotypic features were extracted from published reports using the same questionnaire applied to novel cases.

### Variant Annotation and Scoring

Variants were standardized to the *TECPR2* reference transcript NM_014844.4 (GRCh37/hg19) using Mutalyzer 2.0.32 (Wildeman, van Ophuizen, den Dunnen, & Taschner, 2008) and annotated as described previously (Popp et al., 2017) with up-to-date versions of all tools (Cingolani, Patel et al., 2012; Cingolani, Platts et al., 2012; Freeman, Hart, Gretton, Brookes, & Dalgleish, 2018; Liu, Jian, & Boerwinkle, 2013) and scores (Jian, Boerwinkle, & Liu, 2014; Rentzsch, Witten, Cooper, Shendure, & Kircher, 2019; Xiong et al., 2015) (for details see File S1). All diagnostic *TECPR2* variants were subsequently re-classified (File S3 sheet “TECPR2_variants”) following ACMG guidelines. (Richards et al., 2015)

### Estimation of Carrier Frequencies from Public Databases

We retrieved all *TECPR2* variants from gnomAD (Karczewski et al., 2020) and BRAVO (see Web Resources). These were annotated, scored, and filtered for classification as (likely) pathogenic as described before to calculate carrier frequencies. (Hebebrand et al., 2019)

### Analysis of Missense Variant Spectrum and Modelling of TECPR2 Protein Structure

Distribution of *TECPR2* missense variants in the secondary protein structure was compared to missense variants reported as homozygous in public population databases and protein regions constrained for missense variation were analyzed as described. (Hebebrand et al., 2019) For analysis of the tertiary structure, we used the GalaxyWEB pipeline (Heo, Park, & Seok, 2013; Ko, Park, Heo, & Seok, 2012; Ko, Park, & Seok, 2012) to divide TECPR2 protein sequence into modeling units, predict their structure and refine the top model. Protein data bank (PDB) format structures (Popp & Neuser, 2020) were then used for visualization with a pipeline using the Pymol software (Meyer et al., 2016) and missense clustering analysis as described before. (Hebebrand et al., 2019) For details, also see Supp. Notes S1.

### RNA Expression Analysis for the *TECPR2* variant c.2829del, p.(Asn944Thrfs*7) in P1

Messenger RNA (ribonucleic acid) from peripheral blood lymphocytes of P1 and both parents was used to generate cDNA. Monoallelic expression was analyzed with RT-PCR and Sanger sequencing and *TECPR2* expression was analyzed using quantitative polymerase chain reaction (qPCR) (see details in File S1).

## RESULTS

### TECPR2 Variant Spectrum

Genetic analyses including conventional karyotyping, chromosomal microarray analysis and multi-gene panels (except for P13) were unremarkable in all novel cases. 17 distinct variants in *TECPR2*, including nine truncating and eight missense variants, were identified. Of these, five truncating and five missense variants have not been reported previously (Figure 1A).

**Figure 1.**
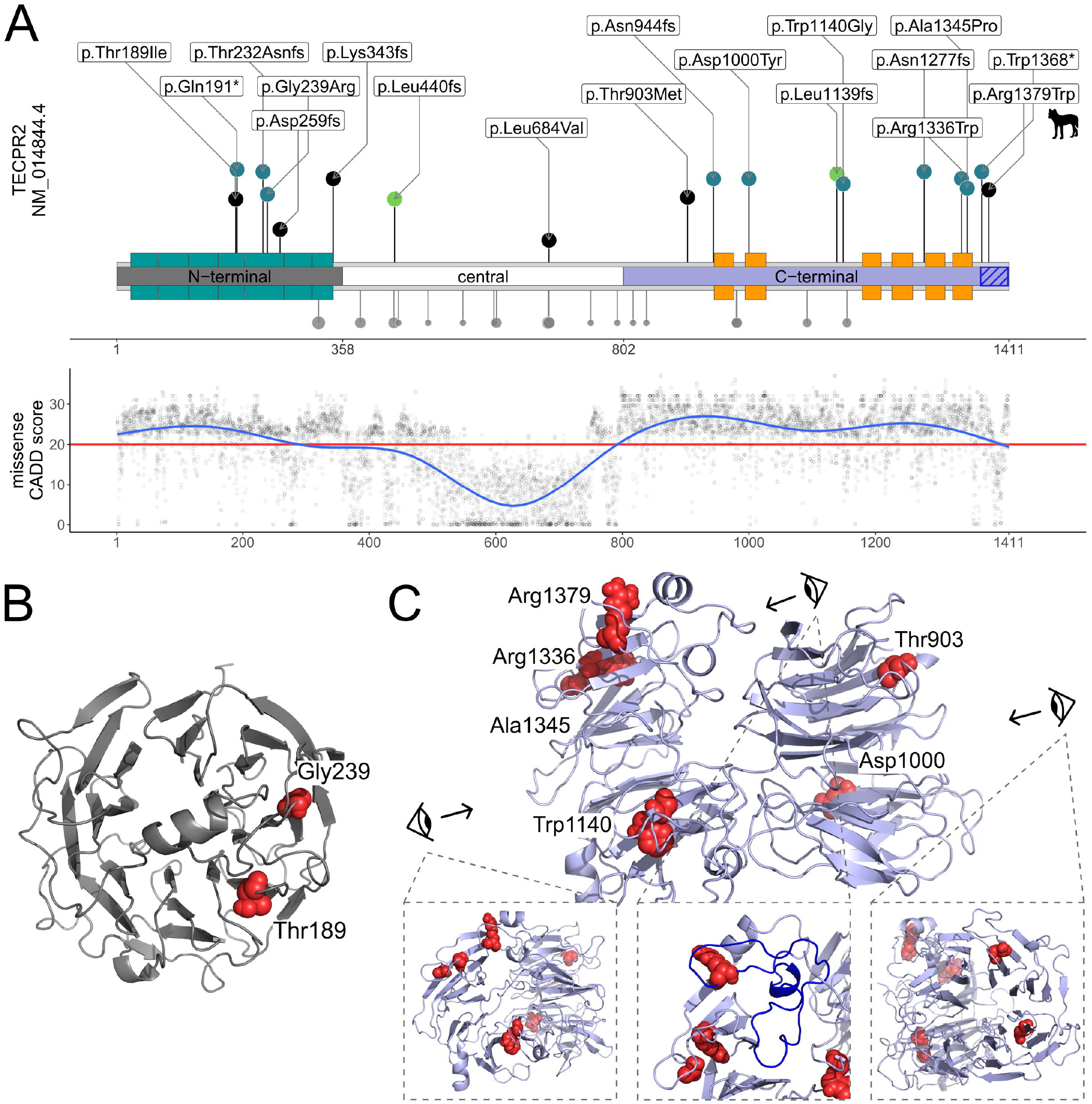
*TECPR2* Structure with Variant Distribution and Computational Scores. (A) Schematic of the *TECPR2* protein with WD40 and TECPR repeat units (WD40: green, TECPR: orange; based on Uniprot O15040) and three modeling units (“N-terminal”: grey, “central”: white, “C-terminal”: purple) identified by GalaxyDom. Disease-associated variants identified in the cohort are depicted towards the top. The length of the segments corresponds to each variants CADD score. Blue dots represent novel identified variants, black dots represent variants reported in the literature and green dots represent the founder variants. Grey dots downwards show homozygous variants from gnomAD, the dot size represents the logarithm of the allele count. In the panel below, a generalized additive model shows the values of CADD PHRED v1.6 for all possible missense variants in *TECPR2* across the protein secondary structure. The red horizontal line marks the recommended cut-off (20). (B) Homology model of the N-terminal domain (AA 1 to 357; grey) generated through the GalaxyTBM pipeline showing the 7-bladed β-propeller fold typical for WD40 repeat. Position of missense variants identified in the individual P3 (Gly239) from our study and “Family E II-1” (Thr189) from the literature review are presented as red spheres. Both missense variants affect conserved residues in β-propeller folds. (C) Lateral overview of the homology model of the C-terminal domain (AA 802 to 1411; blue) showing the two β-propeller folds in the TECPR repeat unit. Position of missense variants identified in the individuals P7 and P8 (Asp1000), P6 (Trp1140), P17 (Arg1336) and P3 (Ala1345) from our study and “Family H I-1” (Thr903) from the literature review and (Arg1379) from the Spanish water dogs (Supp. Notes S1 and Supp. Figure S1) are presented as red spheres. The blue highlighted part of the protein structure in the middle panel is truncated by the most downstream stop gained variant c.4103G>A, p.(Trp1368*) identified in P14 and contains the amino acid position described as pathogenic in Spanish water dogs.

### Founder Variants

The first reported founder variant (Oz-Levi et al., 2012) in the Jewish Bukharian population c.3416del, p.(Leu1139Argfs*75) was identified in the homozygous allelic state in five individuals from the literature and in two cases in our cohort. Additionally, the variant was discovered in compound heterozygous state with the Jewish Ashkenazi founder variant in one previously reported individual. Two previously reported individuals and four cases in our cohort were homozygous for the founder variant in the Jewish Ashkenazi population, c.1319del, p.(Leu440Argfs*19). This variant was also found in compound-heterozygous state with a missense variant(Heimer et al., 2016) and another truncating variant (in our cohort). The two founder variants are located in exons 8 and 16 respectively. GnomAD minor allele frequency (MAF) was 37/275,698 for c.1319del, p.(Leu440Argfs*19) and 2/247,472 for c.3416del, p.(Leu1139Argfs*75). There were no entries for homozygous occurrence of these variants in the reference populations with data available.

### Other Truncating Variants

Among the cases derived from the literature, one individual carried compound heterozygous frameshift variants (c.774del, p.(Asp259Metfs*44); c.1028_1032del, p.(Lys343Argfs*2)). Novel identified truncating variants were c.571C>T, p.(Gln191*) (homozygous), c.694dup, p.(Thr232Asnfs*15) (homozygous), c.2829del, p.(Asn944Thrfs*7) (homozygous), c.3830del, p.(Asn1277Thrfs*43) (compound heterozygous with Ashkenazi founder variant) and c.4103G>A, p.(Trp1368*) (homozygous). The variants are located in exons 5, 6, 7, 12, 18 and 20. MAF in the heterozygous state was consistent with ultrarare variant alleles (Hansen et al., 2019) and between 0 and 2/251,490 (gnomAD).

### Expression analysis of the Stop Codon Containing Transcript in P1

Sanger sequencing of cDNA showed comparable detection of the normal allele and the allele with the c.2829del, p.(Asn944Thrfs*7) variant in both carrier parents of individual P1 (Figure 2A). Additionally, RT-PCR indicated normal expression in individual P1 who is homozygous for the variant (Figure 2B). Comparable expression of *TECPR2* in individual P1, his parents and in-house controls was confirmed by qPCR (Figure S3).

**Figure 2.**
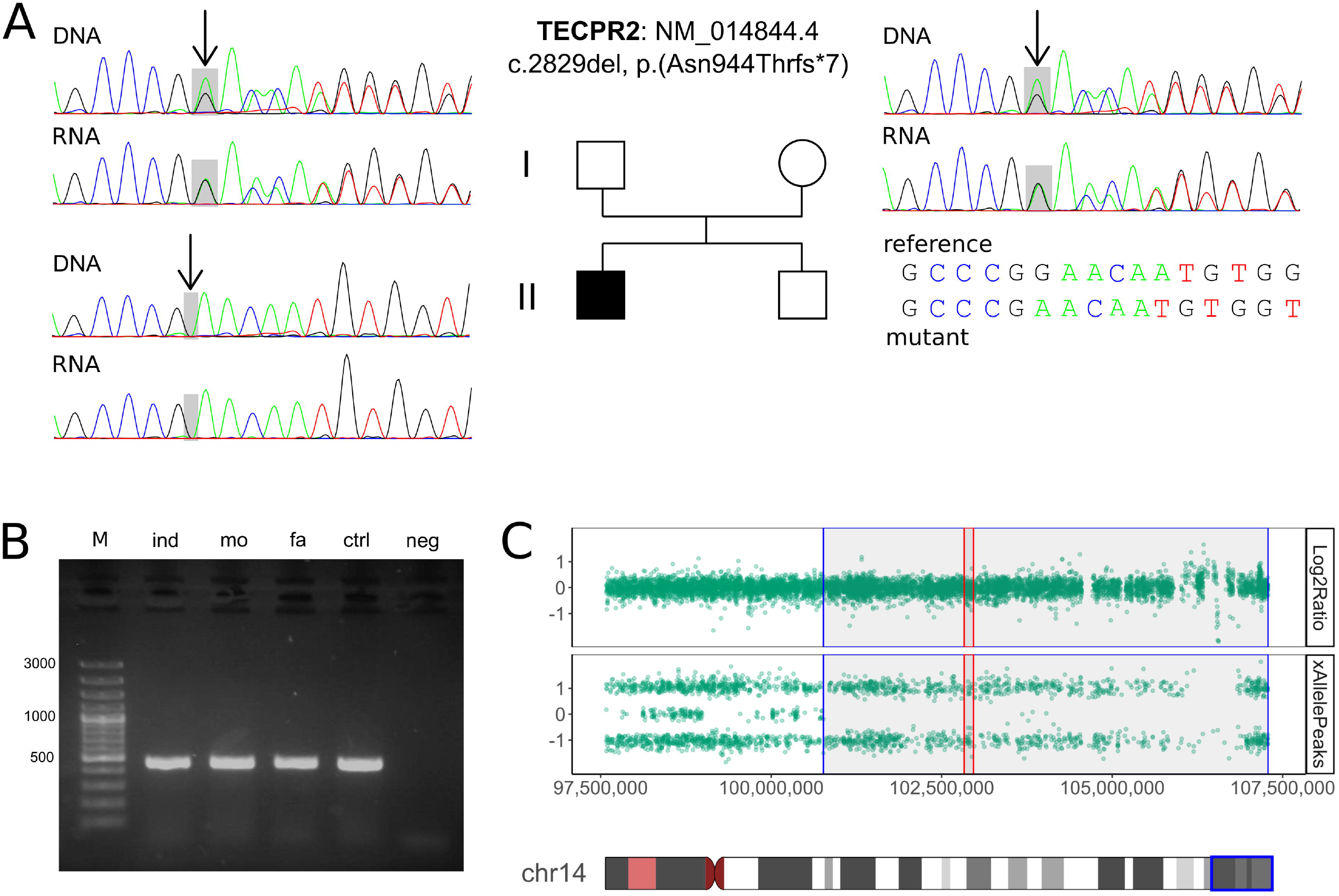
Exemplary Sanger Sequences, RT-PCR and CMA Results for P1. (A) Chromatograms of DNA (Sanger sequencing) and RNA (RT-PCR on PAXgene stabilized blood) of P1 (down left) and his parents (up left and right). (B) Gel electrophoresis of cDNA-amplicon. (C) Chromosomal microarray data for individual P1 showing an unremarkable copy number of chromosome 14 (Log2Ratio top) and SNP allele peak distribution (AllelePeaks bottom) showing a 6.52 Mb run-of-homozygosity (blue) containing *TECPR2* (red).

### Missense Variants

To date, only three disease-associated missense variants have been reported (c.566C>T, p.(Thr189Ile); c.2050C>G, p.(Leu684Val); c.2708C>T, p.(Thr903Met)). Novel variants identified include three homozygous missense variants c.2998G>T, p.(Asp1000Tyr), c.3418T>G, p.(Trp1140Gly) and c.4006C>T, p.(Arg1336Trp) as well as two compound heterozygous missense variants c.715G>A, p.(Gly239Arg) and c.4033G>C, p.(Ala1345Pro). All variants are predicted to be deleterious by multiple *in silico* prediction programs except for the previously described variant c.2050C>G, p.(Leu684Val) (CADD PHRED v1.6: 5.5; mean for all reported missense: 24.4). For a complete overview of *in silico* analyses please refer to File S3. Similar results were obtained for the MAF, which is between 0 and 21/282,852, again except for c.2050C>G, p.(Leu684Val) which showed a MAF of 11,974/282,150. In addition, this variant is found homozygous in gnomAD (440x).

Analysis of spatial distribution in the linear protein structure indicated that missense variants identified in bi-allelic state in individuals with *TECPR2*-associated disease are predominantly located in the N-terminal (amino acid (AA) 1 to 357) and C-terminal (AA 802 to 1,411) protein regions. These two regions display a higher restrain for missense variation as indicated by higher computational scores and depletion of homozygous missense variants (Figure 1A; Figure S1).

This finding is further supported by the missense variant described in Spanish water dogs (Hahn et al., 2015), which is highly conserved (CADD PHRED score v1.6: 27.2) and located near to the c.4033G>C, p.(Ala1345Pro) variant (P3) in the C-terminal region; also the amino acid residue affected by this variant is truncated by the late stop variant c.4103G>A, p.(Trp1368*) identified in P14 (see Supp. Notes S1 and Supp. Figure S1).

Our spatial proximity analysis using predicted 3D protein structures failed to identify clusters of missense variants (Table S3), but showed that all affect highly conserved residues in the repeats forming the N-terminal 7-bladed WD40 β-propeller or the two predicted C-terminal β-propeller structures (Figure 1B and 1C; Figure S2). While we choose the GalaxyTBM (Ko, Park, & Seok, 2012) model for visualization of the spatial missense distribution in Figure S1, the structural similarity of the model predicted *de novo* by the trRosetta algorithm (Yang et al., 2020) is remarkable (Figure S2, Table S2). This convergence of structure prediction algorithms add confidence to the derived models and will thus accelerate our understanding of missense variants in genetic disorders lacking experimentally derived protein structures.

### Carrier Frequency for (Likely) Pathogenic *TECPR2* Variants

Our results indicate that at least 1 in 1,221 individuals (0.082%) in gnomAD and 1 in 1,610 individuals (0.062%) in BRAVO is a carrier of a (likely) pathogenic variant in *TECPR2*. In gnomAD, we were able to estimate the carrier frequency for eight subpopulations, which ranged from 1 in 155 (0.650%; Jewish Ashkenazi) to 1 in 7,654 (0.013%; South Asian). Using these frequencies, the expected incidence is at least 1 in 5,961,640 newborns (based on gnomAD) to 1 in 10,366,419 newborns (based on BRAVO). Of the analyzed populations (which did not include the Jewish Bukharian population) the highest incidence is expected in the Jewish Ashkenazi population with 1 in 95,864 newborns.

### Predicted Tertiary TECPR2 Protein Structure

The three different protein modelling algorithms that we have used (Popp & Neuser, 2020), indicated similar results for the overall TECPR2 tertiary structure. The N-terminal domain (AA 1 to 357) containing seven WD-repeats is predicted to form a 7-bladed β-propeller fold (WD40 domain) with high similarity in all models generated. The central region (AA 358 to 801) could either not be modelled completely due to lack of template structures or resulted in unstructured and highly diverging models. The C-terminal domain (AA 802 to 1,411), containing the six TECPR-repeats annotated from UniProt, was predicted to form a double β-propeller motif in most models with good structural similarity and five to seven blades per propeller. Overall, this indicates a structured C-terminal WD40-domain and TECPR-repeat containing structured double β-propeller motif in the C-terminus, linked by a 444 AA long unstructured peptide (Figure 1 B and C; Figure S2).

### Clinical Spectrum

In our cohort of newly diagnosed cases, 11 of 17 individuals were male. Age at last follow up was between 16 months and 15 years with a mean of 65.2 ± 43.7 (SD) months. Consanguinity was reported in 7 out of the 15 families. Five families were of Jewish Ashkenazi descent, two families were of Jewish Bukharian. Except P1, all individuals were born at term without significant pre-or perinatal complications. Three individuals were small for gestational age. Head circumference at birth was generally within normal limits. At last follow up only seven individuals displayed short stature with a height below −2 SD (standard deviation) from age-matched controls, however, all 11 individuals with data available were below average height. Brachycephaly and microcephaly were observed in seven and four individuals, respectively, with three individuals presenting both. Distinct facial features were seen in 11 individuals though were not uniform. Shared characteristics included a short neck, synophrys and a triangular-shaped face, a recognizable pattern, or facial gestalt, was not appreciated. Skeletal abnormalities including significant lumbar kyphosis, a barrel-shaped chest or hyperextension of the neck were present in five cases.

The ages at diagnosis in our cohort ranged between 13 months and 15 years with a mean of 55.6 ± 48.8 (SD) months. All affected individuals showed global developmental delay and later intellectual disability (DD/ID) in the mild (n=1), moderate (n=7) and severe (n=8) ranges. P2 had only mildly delayed gross motor skills at last investigation, but her young age rendered a detailed assessment difficult. Six individuals with moderate or severe development delay were reported to have behavioral dysregulation with hyperactivity, restlessness, and aggressive behaviors. Two received a formal diagnosis of autism spectrum disorder. Ten children (age range: 16 months to 8 years) had not started walking at the time of last follow up and 7 individuals walked independently (mean age: 40.5 ± 36.2 (SD) months). P3 was diagnosed with dystonic/dyskinetic cerebral palsy and started walking around the age of 10 years. Speech development was delayed in all children and speech remained limited to a few words with five individuals remaining completely non-verbal.

Most common neurological manifestations in our cohort included axial and appendicular hypotonia (17/17) accompanied by gait ataxia (11/11), hyporeflexia of the lower limbs (13/17) and dysarthria (6/8). Autonomic dysfunction, e.g. temperature instability (3/14), and hyperhidrosis (2/14) were noticed in a subset of cases (5/15). Four individuals were reported to have impaired pain sensation (4/16). Febrile seizures were found in P1 as well as P10; P13, P14 and P15 were reported to have medically-refractory epilepsy and a peripheral neuropathy was diagnosed in P6. Hearing impairment (3/13) and visual impairment (5/12) were present in a subset. The constellation of central respiratory dysregulation, dysphagia, and neuromuscular-derived respiratory insufficiency was common, resulting in central nocturnal (8/13) and/or daytime (5/16) hypoventilation, dysphagia (9/17) and impaired clearance of secretions. This was complicated by recurrent respiratory infections (14/15), aspiration events (10/15), gastroesophageal reflux disease (9/15) and necessitated non-invasive positive pressure ventilation (i.e. nocturnal BiPAP) (2/13), and utilization of gastrostomy tubes (6/11) in a subset. Airway malformation such as laryngeal cleft or laryngomalacia were identified in a subset (4/17). Five individuals (5/15) were reported to have chronic and significant constipation.

Clinical manifestations of previously reported individuals are summarized in File S2. One case (Covone et al., 2016) was excluded from further analysis since the variant c.2050C>G, p.(Leu684Val) was classified as likely benign according to ACMG criteria. In summary, manifestations shared by the majority of all 27 individuals include: Global development delay and intellectual disability (26/26, 100%), muscular hypotonia (27/27, 100%), hyporeflexia of the lower limbs (22/27, 83%) and gait ataxia (19/19, 100%). Peripheral neuropathy, dysarthria and abnormal facial features were found in 9/12 (75%), 12/14 (86%) and 19/25 (76%) of individuals with sufficient data available. Recurrent respiratory infections (23/25, 92%), gastroesophageal reflux in infancy (18/25, 72%) and nocturnal hypoventilation (12/17, 71%) affected most individuals.

### Brain Imaging and EEG

Review of 16 brain MRI studies from our cohort (Figure 3 and Figure S4) and a review of reported cases in the literature defined a thin corpus callosum as a common feature (11/21, 52%). Additional findings in a subset of individuals included mild ventriculomegaly (often asymmetric colpocephaly), delayed myelination and diffuse cerebral atrophy. EEG (electroencephalogram) was abnormal in four cases (4/15, 27%), but no specific pattern was reported.

**Figure 3.**
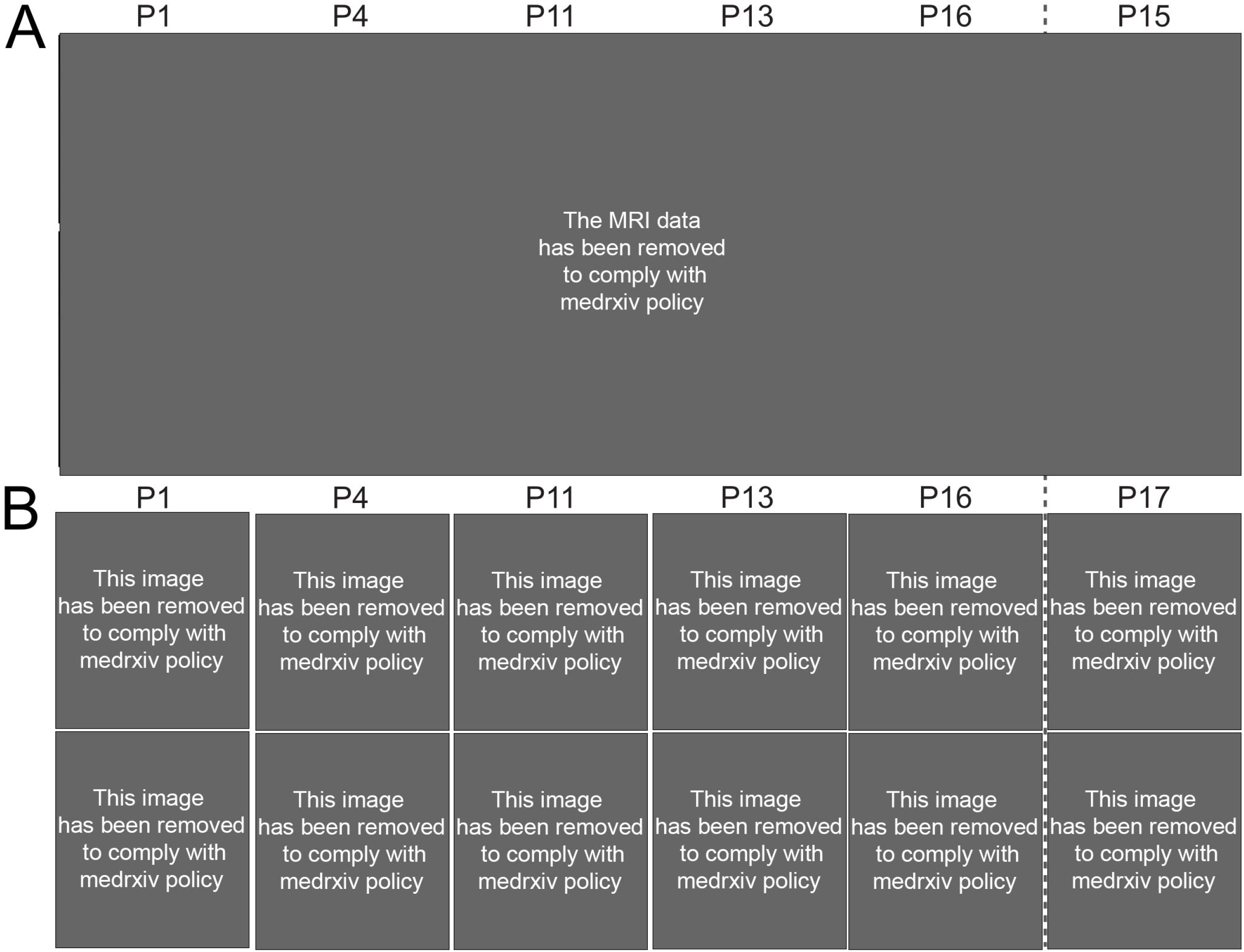
MRI and Facial Features of Individuals with *TECPR2*-associated Disease. (A) MRI data removed from the preprint version to comply with medrxiv policy. (B) Facial images removed from the preprint version to comply with medrxiv policy.

## DISCUSSION

We here report a series of 17 individuals with bi-allelic *TECPR2* variants from eight non-consanguineous families and nine consanguineous families, and combine the detailed clinical, imaging, and molecular characterization of these individuals with the 11 cases previously reported. Since the variant c.2050C>G, p.(Leu684Val) was classified as likely benign according to ACMG criteria, one previously reported case (Covone et al., 2016) was excluded. The girl’s different clinical presentation without developmental delay, autonomic nervous system involvement or abnormal facial shape supports the variant assessment. Additionally, an inherited variant of unknown significance in *SPG7* was reported as an additional genetic finding (Covone et al., 2016). The analysis of the remaining 27 individuals defines a core set of clinical and molecular features. These consist of global developmental delay and intellectual disability, axial and appendicular hypotonia, dysarthria and an abnormal gait, often described as an ataxic gait. Peripheral neuropathy was found in two thirds of all individuals in whom a detailed neurological assessment was available. Along with this, hyporeflexia was common and signs of autonomic dysfunction were prominent in the majority of cases. The latter included central hypoventilation, impaired temperature, and blood pressure regulation, repeat aspiration events and evidence of abnormal gastrointestinal motility. These features imply involvement of both the central and peripheral nervous systems and substantiate features of hereditary sensory and autonomic neuropathy (HSAN).

Whereas spasticity was recognized as a hallmark feature in the individuals initially reported (Oz-Levi et al., 2012), the overall prevalence of spasticity was limited to a subset in our analysis (24%). We recognize that this is a potentially age-dependent manifestation, since increased tone was mainly reported in older individuals. P3 stands out because of the presence of dystonia, which was not present in previously published cases and possibly further broadens the spectrum of neurological symptoms. Of note, epilepsy was relatively infrequent in our cohort and consisted of two individuals who experienced febrile seizures, two previously reported siblings with infrequent generalized tonic-clonic seizures and three individuals with medically refractory seizures. Future studies will be necessary to reassess epilepsy as an associated feature. Overall, the wide neurological manifestations in individuals with *TECPR2*-associated disease along the age spectrum, point to an involvement of multiple areas of the central nervous system (i.e. cortico-spinal tracts, cerebral cortex, brain stem, possibly basal ganglia) as well as the peripheral nervous system. A large part of the morbidity and mortality associated with *TECPR2* results from central hypoventilation requiring therapy with non-invasive positive pressure ventilation and occasionally active mechanical ventilatory support. Our findings are supported by a recently published, detailed analysis of the distinct breathing pattern from one affected individual. (Patwari et al., 2020)

Based on our clinical experience and the reported disease manifestations, we suggest a framework for routine surveillance as detailed in Table 2. Symptomatic treatment should be tailored to each individual case and aims at preserving function and preventing long-term morbidity and mortality. Early developmental support should be maximized to harness the developmental potential.

Overall, our cross-sectional analysis suggests that there is evidence of disease progression from a predominantly neurodevelopmental disorder with global developmental delay and hypotonia in early childhood to a progressive disease with corticospinal and corticobulbar dysfunction later in life. We know from personal communications about the disease course of previously reported patients (Heimer et al., 2016; Oz-Levi et al., 2012), who all lost the ability to walk.

Due to largely non-specific initial clinical features, individuals with *TECPR2*-associated disease may initially receive a diagnosis of cerebral palsy. In addition to an often-unremarkable perinatal history, clinical features that help distinguish *TECPR2*-disease from cerebral palsy include the findings of central apnea/hypoventilation, autonomic instability, hyporeflexia as well as other signs of peripheral neuropathy. Brain MRI in *TECPR2*-associated disease shows a thinning of the posterior parts of the corpus callosum in about half of individuals. This finding can help guide diagnostic testing.

A diagnosis is achieved through molecular testing. With the identification of novel truncating and missense variants we confirm and broaden the spectrum of disease-associated variants in *TECPR2*-associated hereditary sensory and autonomic neuropathy with intellectual disability. All individuals in the cohort with distinct ethnic origin carried the respective founder variant. This observation affirms the expected genotypic trait in the Jewish Ashkenazi and Jewish Bukharian population. However, the identification of other truncating variants provides evidence for the occurrence of *TECPR2*-associated disease in other ethnic groups. For all families with homozygous variants other than the founder variants, consanguinity of the parents was reported. This is exemplified for P1 where the run-of-homozygosity on chromosome 14 was not described in the CMA report, because it was below the 10 Mb (mega base) filtering cutoff (Figure 2C). Similar results were reported for P2 and P16 (File S1).

Our analysis did not show clustering or specific distribution pattern of the truncating variants. RNA analysis of the novel frameshift variant c.2829del, p.(Asn944Thrfs*7), identified in P1, indicated escape from nonsense-mediated decay. This argues against NMD and is in line with previous results in cell-lines showing no effect on mRNA levels for the Jewish Bukharian variant c.3416del, p.(Leu1139Argfs*75) (Oz-Levi et al., 2012), but instead the resulting truncated protein being targeted for proteasome mediated degradation after translation.

In contrast, all disease-associated missense variants in this cohort affect conserved residues in repeats forming the blades of β-propeller structures at the C-terminal and the N-terminal ends of the protein (Figure 1). As we could not identify clustering in the tertiary structure, misfolding and subsequent degradation could cause loss of the protein carrying these missense substitutions. All five individuals from our cohort harboring missense variants showed moderate to severe DD/ID and are as severely affected as individuals with truncating variants. Therefore, our data do not indicate milder clinical manifestations in carriers of missense variants. This clinical observation further supports a similar pathomechanism, e.g. degradation of truncated or misfolded proteins, for both truncating and missense variants. However, due to the currently limited knowledge about *TECPR2* function and lack of well-established and readily available functional tests, in most cases missense variants cannot be classified as (likely) pathogenic according to ACMG guidelines. Based on our computational analyses, we propose to consider the following criteria for the interpretation of *TECPR2* missense variants: 1) variant position in the functional domains identified through our conservation and modelling analyses (PM1_Supporting; Figure 1A; Figure S1 and S2), 2) deleterious effect predicted by *in silico* CADD score with cutoff >20 (PP3; Figure 1A), 3) the patient’s phenotype matches the core features as well as *TECPR2*-specific symptoms of the HSAN-spectrum (Table 1) and exome wide analyses does not reveal other clinically relevant findings (PP4), 4) co-segregation of the identified variants with multiple affected family members (PP1).

**Table 1.** Clinical Manifestations of *TECPR2*-associated Disease.

| Group | Phenotype | HPO | Novel cases (%) |  | Literature cases (%) |  | All cases (%) |  |
| --- | --- | --- | --- | --- | --- | --- | --- | --- |
| Phenotypical abnormalities of body and face | Abnormal facial shape | HP:0001999 | 69 | (11/16) | 89 | (8/9) | **76** | (19/25) |
|  | Short stature | HP:0004322 | 44 | (7/16) | 78 | (7/9) | 56 | (14/25) |
|  | Abnormality of skeletal morphology | HP:0011842 | 29 | (5/17) | 100 | (2/2) | 37 | (7/19) |
|  | Microcephaly | HP:0000252 | 47 | (7/15) | 75 | (6/8) | 57 | (13/23) |
|  | Brachycephaly | HP:0000248 | 25 | (4/16) | 56 | (5/9) | 36 | (9/25) |
| Intellectual and social development | Behavioral abnormalities | HP:0000708 | 43 | (6/14) | 100 | (4/4) | 56 | (10/18) |
|  | Global developmental delay; Intellectual disability | HP:0001263;<br>HP:0001249 | 100 | (16/16) | 100 | (10/10) | **100** | (26/26) |
|  | *moderate* | HP:0011343;<br>HP:0002342 | 44 | (7/16) | 14 | (1/7) | 35 | (8/23) |
|  | *severe* | HP:0011344;<br>HP:0010864 | 50 | (8/16) | 86 | (6/7) | 61 | (14/23) |
| Neurological system | Muscular hypotonia | HP:0001252 | 100 | (17/17) | 100 | (10/10) | **100** | (27/27) |
|  | Hyporeflexia of lower limbs | HP:0002600 | 76 | (13/17) | 90 | (9/10) | **81** | (22/27) |
|  | Gait ataxia | HP:0002066 | 100 | (11/11) | 100 | (8/8) | **100** | (19/19) |
|  | Dysarthria | HP:0001260 | 75 | (6/8) | 100 | (6/6) | **86** | (12/14) |
|  | Peripheral neuropathy | HP:0009830 | 25 | (1/4) | 100 | (8/8) | **75** | (9/12) |
|  | Impaired pain sensation | HP:0007328 | 25 | (4/16) | 100 | (4/4) | 40 | (8/20) |
|  | Abnormality of the autonomic nervous system | HP:0002270 | 33 | (5/15) | 100 | (10/10) | 60 | (15/25) |
|  | Abnormal systemic blood pressure | HP:0030972 | 0 | (0/13) | 88 | (7/8) | 33 | (7/21) |
|  | Visual impairment | HP:0000505 | 44 | (7/16) | 75 | (3/4) | 50 | (10/20) |
|  | Hearing impairment | HP:0000365 | 24 | (4/17) | 100 | (2/2) | 32 | (6/19) |
| Respiratory system | Recurrent respiratory infections | HP:0002205 | 93 | (14/15) | 90 | (9/10) | **92** | (23/25) |
|  | Aspiration | HP:0002835 | 67 | (10/15) | 50 | (4/8) | 61 | (14/23) |
|  | Nocturnal hypoventilation | HP:0002877 | 62 | (8/13) | 100 | (4/4) | **71** | (12/17) |
|  | Central hypoventilation | HP:0007110 | 31 | (5/16) | 100 | (9/9) | 56 | (14/25) |
| GI system | Gastroesophageal reflux at infancy | HP:0002020 | 60 | (9/15) | 90 | (9/10) | **72** | (18/25) |
|  | Dysphagia | HP:0002015 | 53 | (9/17) | 11 | (1/9) | 38 | (10/26) |
| diagnostic procedures | Abnormal corpus callosum morphology | HP:0001273 | 53 | (8/15) | 50 | (3/6) | 52 | (11/21) |
|  | Abnormality of metabolism/homeostasis | HP:0001939 | 62 | (8/13) | 25 | (1/4) | 53 | (9/17) |

**Table 2.**
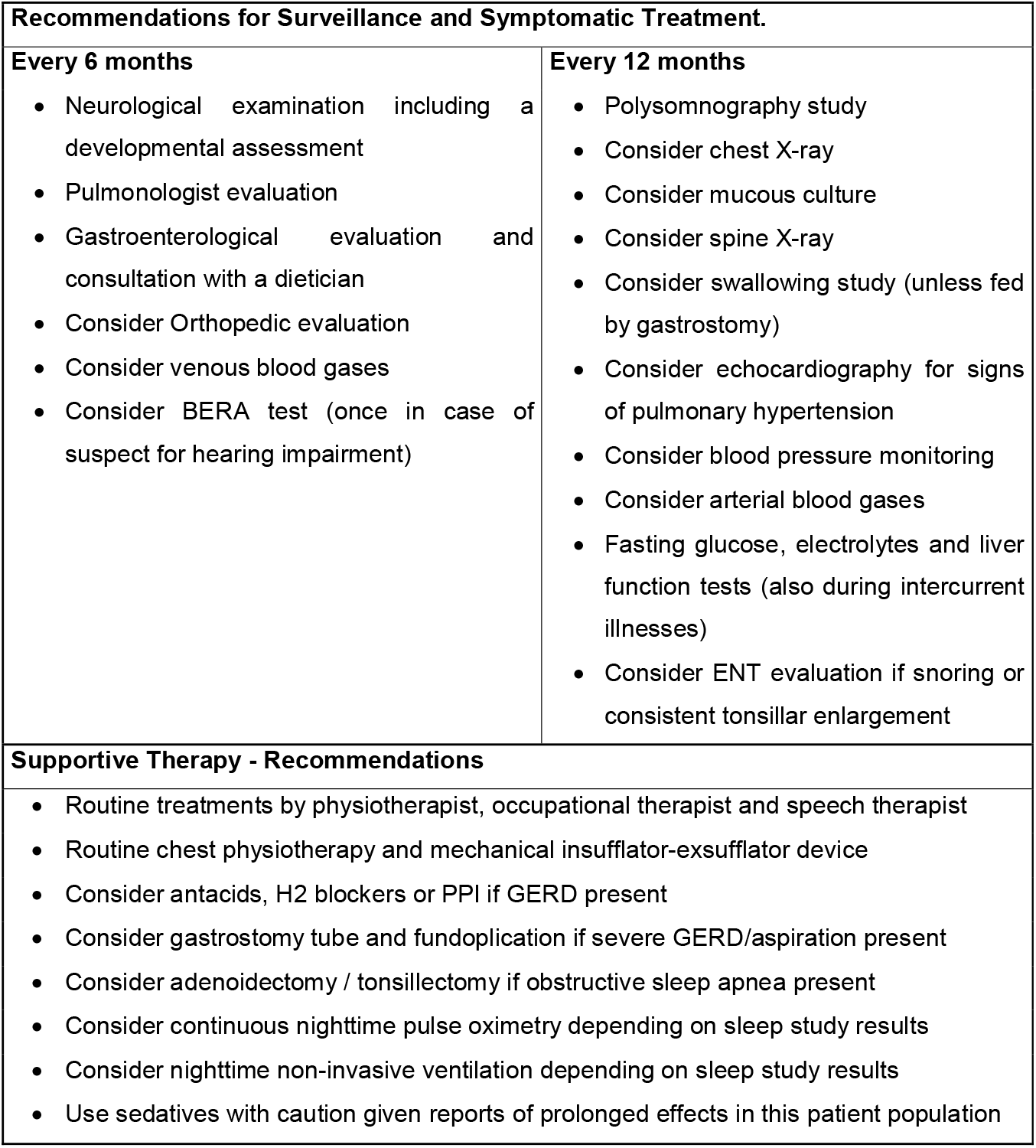
Recommendations for Surveillance and Symptomatic Treatment.

| **Recommendations for Surveillance and Symptomatic Treatment.** |  |
| --- | --- |
| **Every 6 months**<br>• Neurological examination including a developmental assessment<br>• Pulmonologist evaluation<br>• Gastroenterological evaluation and consultation with a dietician<br>• Consider Orthopedic evaluation<br>• Consider venous blood gases<br>• Consider BERA test (once in case of suspect for hearing impairment) | **Every 12 months**<br>• Polysomnography study<br>• Consider chest X-ray<br>• Consider mucous culture<br>• Consider spine X-ray<br>• Consider swallowing study (unless fed by gastrostomy)<br>• Consider echocardiography for signs of pulmonary hypertension<br>• Consider blood pressure monitoring<br>• Consider arterial blood gases<br>• Fasting glucose, electrolytes and liver function tests (also during intercurrent illnesses)<br>• Consider ENT evaluation if snoring or consistent tonsillar enlargement |
| **Supportive Therapy - Recommendations** |  |
| • Routine treatments by physiotherapist, occupational therapist and speech therapist<br>• Routine chest physiotherapy and mechanical insufflator-exsufflator device<br>• Consider antacids, H2 blockers or PPI if GERD present<br>• Consider gastrostomy tube and fundoplication if severe GERD/aspiration present<br>• Consider adenoidectomy / tonsillectomy if obstructive sleep apnea present<br>• Consider continuous nighttime pulse oximetry depending on sleep study results<br>• Consider nighttime non-invasive ventilation depending on sleep study results<br>• Use sedatives with caution given reports of prolonged effects in this patient population |  |

Our estimation of carrier frequency is based on automated ACMG classification of variants and therefore includes only potentially truncating variants. Given that eight of the 17 unique variants are missense variants and considering the large number of uncharacterized *TECPR2* missense variants (729 in gnomAD and 448 in BRAVO), we anticipate that the true carrier frequency for (likely) pathogenic *TECPR2*-variant might be double our current estimate of 0.082% in the general population. Notably, the estimated carrier frequency is 7.9x higher (0.650%) in individuals with Jewish Ashkenazi background and at least 16.2x higher (1.33%) with Jewish Bukharian background. (Oz-Levi et al., 2012) A review of carrier screening tests for individuals of Jewish descent showed that *TECPR2* is currently included in four offered tests (see Table S1). Overall, based on these high carrier frequencies, both founder variants should be included in commercial carrier screening tests to inform genetic counseling and diagnostics in Jewish couples at increased risk for children with *TECPR2*-associated disease.

*TECPR2* encodes a protein that is implicated in the early steps of the autophagy pathway where it interacts with the Atg8 family proteins, including LC3, to promote autophagic vesicle formation. (Behrends, Sowa, Gygi, & Harper, 2010) Fibroblasts from affected individuals showed a decreased number of autophagosomes and reduced delivery of LC3 and p62 for lysosomal degradation; this suggests an impairment of autophagic flux. (Oz-Levi et al., 2012) Providing insights into the mechanism of defective autophagy, a subsequent study showed that TECPR2 is involved in maintaining functional endoplasmic reticulum exit sites, which are implicated in the cargo from endoplasmic reticulum to Golgi and may serve as scaffolds for the formation of autophagosomes. (Stadel et al., 2015)

While the precise role of autophagy in *TECPR2*-associated disease remains to be established, there are several clinical features that are shared with other single gene disorders of this pathway. (Ebrahimi-Fakhari et al., 2016; Teinert et al., 2019) This includes the involvement of multiple brain areas, clinical signs that point to a progressive involvement of the long central nervous system tracts, such as the cortico-spinal tracts, as well as the imaging finding of a thinning of the corpus callosum. *TECPR2*-associated disease, however, stands out for its prominent involvement of brain stem function, autonomic dysregulation, and peripheral neuropathy.

In summary, our cross-sectional analysis provides a depiction of clinical and molecular features across the age spectrum. Functional analyses of the variant mechanisms are of great importance to confirm the intended effect by our *in silico* modelling approach. Future prospective longitudinal studies to better define the natural history and patterns of disease progression. Our present study provides a framework for assessing disease manifestations. Close follow up and surveillance for neurological and non-neurological manifestations is recommended.

## Data Availability

All data generated or analyzed during this study can be found in the supplementary materials provided. Some of the data had to be removed from the preprint version to comply with medrxiv policy.

https://doi.org/10.5281/zenodo.4050472

## ACKNOWLEDGEMENTS

We thank all involved families for participating in this study. We also thank the Exome Aggregation Consortium and therein involved groups for providing exome and genome variant data for comparison. B.B. is supported by scholarships from the German National Academic Foundation and the Carl Duisberg Program by the Bayer Foundation. B.P. is supported by the Deutsche Forschungsgemeinschaft (DFG) through grant PO2366/2–1. D.E-F. is supported by grants from the CureAP4 Foundation, CureSPG50 Foundation, Spastic Paraplegia Foundation, the Thrasher Research Fund, and a joint research agreement with Astellas Pharmaceutical Inc. and MitoBridge Inc.. Trio-ES of P17 was supported by the US NIH National Human Genome Research Institute (NHGRI)/National Heart Lung and Blood Institute (NHLBI) UM1 HG006542 (J.R.L.). J.E.P. is supported by NHGRI K08 HG008986. This research was made possible through access to the data and findings generated by the 100,000 Genomes Project. The 100,000 Genomes Project is managed by Genomics England Limited (a wholly owned company of the Department of Health and Social Care). The 100,000 Genomes Project is funded by the National Institute for Health Research and NHS England. The Wellcome Trust, Cancer Research UK and the Medical Research Council have also funded research infrastructure. The 100,000 Genomes Project uses data provided by patients and collected by the National Health Service as part of their care and support. Funding for this study was provided by the Baylor-Hopkins Center for Mendelian Genomics through National Human Genome Research Institute grant 5U54HG006542.

## AVAILABILITY OF DATA AND MATERIALS

All data generated or analyzed during this study can be found in the online version of this article at the publisher’s website. The genomes from P16 and her parents are available within the Genomics England research environment, access to which can be obtained by applying to join a Genomics England Clinical Interpretation Partnership domain (www.genomicsengland.co.uk/join-a-gecip-domain).

## AUTHORS’ CONTRIBUTIONS

B.P., R.A.J. and S.N. conceived the initial study concept and coordinated collection of clinical and genetic data through matchmaking and personal communications. S.N., B.Br. and G.H. reviewed literature data and standardized the clinical HPO terms. A.Se., I.B., L.O., D.S., D.A.S., B.By., W-H.T., B.T.D., S.Sr., P.J., R.J.G., M.Z., Y.D., V.M., J.W., G.E., A.A.T., O.I., A.E.A.N., K.M., F.S.A., H.M.D., M.O.H., B.E.B., Y.H., A.Sz., S.I., F.K., S.E., R.M., H.H., V.S.R., U.K., A.T.P., I.H., J.E.P., J.R.L., C.P., T.M., G.M.R., B.B.-Z., G.H. and D.E.F. provided clinical and genetic data and performed clinical assessments. S.N., I.B., L.O., S.Sr., W.-H.T., G.E., F.S.A., A.E.A.N., H.M.D., M.Z., B.B.-Z., G.H., S.E. and V.S.R. wrote the case reports. I.B. provided RNA samples of P1 and his parents. S.Sc. conducted RT-PCR and qPCR analyses. C.B. performed protein analysis. J.M. supported computational protein modelling. M.K. processed samples of P1 and his parents for establishing a cell line. S.N. and B.P. created Figures 1, 2 and 3 and the Supplementary materials. S.N., B.B., G.H., B.B-Z., D.E-F. and B.P. wrote and edited the manuscript. All authors reviewed and commented the final draft manuscript.

## WEB RESOURCES

gnomAD browser: http://gnomad.broadinstitute.org/

BRAVO/TOPmed: https://bravo.sph.umich.edu

Mutalyzer: https://mutalyzer.nl

ClinVar: https://www.ncbi.nlm.nih.gov/clinvar/

UCSC BLAT: https://genome.ucsc.edu/cgi-bin/hgBlat

UCSC Browser: https://genome.ucsc.edu/cgi-bin/hgGateway

SPIDEX/SPANR: http://tools.genes.toronto.edu

dbscSNV: http://www.liulab.science/dbscsnv.html

## SUPPPLEMENTARY FILES

File S1: clinical reports, supplementary methods, results, figures and tables, references

File S2: comprehensive clinical data

File S3: comprehensive genetic data

## Notes

### Competing Interest Statement

The authors have declared no competing interest.

### Author Declarations

The study adheres to the principles set out in the Declaration of Helsinki. The following Research Ethics Committee approved genetic testing in research setting within the study: Ethical Committee of the Medical Faculty, Leipzig University (P1), Institutional Review Board at Boston Childrens Hospital (IRB-P00033016; P2, P4 and P5), Ethics Review Board of Technical University of Munich (P3), Institutional Review Board of King Faisal Specialist Hospital and Research Center (KFSRHC RAC# 2080006 and 2121053; P7, P8 and P13), Institutional Review Board at University College London (P14 and P15, SYNaPS cohort), East of England and South Cambridge Research Ethics Committee (REC: 14/EE/1112) for 100,00 Genomes Project Protocol (P16), Institutional Review Board at Baylor College of Medicine (H-29697) and Comite Etico Cientifico at Facultad de Medicina, Clinica Alemana Universidad del Desarrollo (P17). Genetic testing for P6, P9, P10, P11 and P12 was performed in a diagnostic setting. The authors received and archived written consent of the legal guardians to publish genetic and clinical data (P1 - P17) as well as photographs, computed tomography (CT) scan and magnetic resonance imaging (MRI) images (P1, P4, P6, P11, P13, P15, P16, P17).

### Summary of Updates

Our preprint posted at medRxiv has in the meantime attracted some attention from clinicians who have seen further individuals with suspected TECPR2-associated disease. Thus, we were able to collect four additional individuals from three families with biallelic TECPR2 variants. We have extended our detailed clinical assessment including the phenotypic descriptions of these individual and our computational anal-yses to the novel variants identified in these families. While the clinical spectrum remains in line with the previous cohort, the novel variants further expand the mutational spectrum adding both a late truncating variant and additional missense variants, strengthening our point that missense variation in TECPR2 likely contributes up to half of the pathogenic variants. Fourteen new coauthors who contributed clinical and genetic data have been added to the revised manuscript version. Taken together, our revised manuscript improves upon our previous comprehensive summary of clinical and molecular data will be the reference for all further reports of TECPR2-associated hereditary sensory and autonomic neuropathy with intellectual disability.

