## Supplementary material for "Clinical, neuroimaging and molecular spectrum of *TECPR2*-associated hereditary sensory and autonomic neuropathy with intellectual disability": File S1

[5] Sackler Faculty of Medicine, Tel Aviv University, Tel Aviv, Israel

[11] kbo-Kinderzentrum München, Munich, Germany

[12] Lehrstuhl für Neurogenetik, Technische Universität München, Munich, Germany

[13] Munich Cluster for Systems Neurology (Synergy), Ludwig-Maximilians-Universität München, Munich, Germany

[14] Department of Neurology, Boston Children’s Hospital, Harvard Medical School, Boston, USA

[15] Department of Anesthesia, Critical Care and Pain Medicine, Boston Children’s Hospital, Harvard Medical School, Boston, USA

[16] Nicklaus Children's Hospital, Miami, USA

[17] Powell Gene Therapy Center, University of Florida, Gainesville, USA

[18] Cincinnati Children’s Hospital Medical Center and the University of Cincinnati College of Medicine, Cincinnati, USA

[19] Department of Neuroscience, King Faisal Specialist Hospital and Research Center, Riyadh, Saudi Arabia

[20] Department of Translational Genomics, Center for Genomic Medicine, King Faisal Specialist Hospital and Research Center, Riyadh, Saudi Arabia

[21] Sheikh Khalifa Medical City, Abu Dhabi, United Arab Emirates

[22] Department of Paediatrics and Child Health, Aga Khan University Hospital, Karachi, Pakistan

[23] Department of Neuromuscular Disorders, Queen Square Institute of Neurology, University College London, London, UK

[24] Oxford Centre for Genomic Medicine, Oxford, UK

[25] NIHR Biomedical Research Centre, Wellcome Centre for Human Genetics, University of Oxford, Oxford, UK

[26] Department of Molecular and Human Genetics, Baylor College of Medicine, Houston, USA

[27] Department of Pediatrics, Baylor College of Medicine, Houston, USA

[28] Texas Children's Hospital, Houston, Texas, USA

[29] Human Genome Sequencing Center, Baylor College of Medicine, Houston, USA

[30] Division of Genetics and Genomics, Boston Children's Hospital, Boston, USA

[31] Section of Pediatric Neurology and Developmental Neuroscience, Department of Pediatrics, Baylor College of Medicine, Houston, USA

[32] Facultad de Medicina, Clinica Alemana Universidad del Desarrollo, Santiago, Chile

[33] Department of Diagnostic and Interventional Radiology, Heidelberg University Hospital, Heidelberg, Germany

[34] Institute of Human Genetics, Friedrich-Alexander-Universität (FAU), Erlangen, Germany

[35] Department of Chemistry, Vanderbilt University, Nashville, USA

[36] Institute for Drug Discovery, University of Leipzig Medical Center, Leipzig, Germany

[37] Department of Anatomy and Cell Biology, College of Medicine, Alfaisal University, Riyadh, Saudi Arabia

[*,#] authors contributed equally to this work

Correspondence to:

Corresponding author: Sonja Neuser, MD

Corresponding author's address: Philipp-Rosenthal-Straße 55

Institute of Human Genetics

University of Leipzig Medical Center

04103 Leipzig, Germany

### DETAILED INDIVIDUAL CASE REPORTS

#### Individual P1

*This case report has been removed from the preprint version to comply with medrxiv policy. Please see the published version or contact the authors if you are interested in this information.*

#### Individual P2

*This case report has been removed from the preprint version to comply with medrxiv policy. Please see the published version or contact the authors if you are interested in this information.*

#### Individual P3

*This case report has been removed from the preprint version to comply with medrxiv policy. Please see the published version or contact the authors if you are interested in this information.*

#### Individual P4

*This case report has been removed from the preprint version to comply with medrxiv policy. Please see the published version or contact the authors if you are interested in this information.*

#### Individual P5

*This case report has been removed from the preprint version to comply with medrxiv policy. Please see the published version or contact the authors if you are interested in this information.*

#### Individual P6

*This case report has been removed from the preprint version to comply with medrxiv policy. Please see the published version or contact the authors if you are interested in this information.*

#### Individual P7

*This case report has been removed from the preprint version to comply with medrxiv policy. Please see the published version or contact the authors if you are interested in this information.*

#### Individual P8

*This case report has been removed from the preprint version to comply with medrxiv policy. Please see the published version or contact the authors if you are interested in this information.*

#### Individual P9

*This case report has been removed from the preprint version to comply with medrxiv policy. Please see the published version or contact the authors if you are interested in this information.*

Individual P10

*This case report has been removed from the preprint version to comply with medrxiv policy. Please see the published version or contact the authors if you are interested in this information.*

Individual P11

*This case report has been removed from the preprint version to comply with medrxiv policy. Please see the published version or contact the authors if you are interested in this information.*

Individual P12

*This case report has been removed from the preprint version to comply with medrxiv policy. Please see the published version or contact the authors if you are interested in this information.*

Individual P13

*This case report has been removed from the preprint version to comply with medrxiv policy. Please see the published version or contact the authors if you are interested in this information.*

Individual P14

*This case report has been removed from the preprint version to comply with medrxiv policy. Please see the published version or contact the authors if you are interested in this information.*

Individual P15

*This case report has been removed from the preprint version to comply with medrxiv policy. Please see the published version or contact the authors if you are interested in this information.*

Individual P16

*This case report has been removed from the preprint version to comply with medrxiv policy. Please see the published version or contact the authors if you are interested in this information.*

Individual P17

*This case report has been removed from the preprint version to comply with medrxiv policy. Please see the published version or contact the authors if you are interested in this information.*

### SUPPLEMENTARY METHODS

#### Variant Annotation

Variants were converted to VCF format using VariantValidator (version 1.0.4.dev47+gfd41f45) (Freeman, Hart, Gretton, Brookes, & Dalgleish, 2018) and annotated with scores from the dbNSFP database (version 2.93) (Liu, Jian, & Boerwinkle, 2013) using SnpEff/SnpSift (v4.3.1t). (Cingolani, Patel et al., 2012; Cingolani, Platts et al., 2012) CADD score values (version 1.6) (Rentzsch, Witten, Cooper, Shendure, & Kircher, 2019) were annotated through the online "score" functionality using the VCF as input. For splicing prediction, SPIDEX/SPANR (Xiong et al., 2015) and dbscSNV scores (Jian, Boerwinkle, & Liu, 2014) were annotated using SnpSift and the files provided from the respective website.

#### Estimation of Carrier Frequencies from Public Databases

*TECPR2* variants were downloaded from gnomAD (v2.1.1 assessed on July 27$^{th}$, 2020) (Karczewski et al., 2020) and BRAVO (freeze5 assessed on July 27$^{th}$, 2020) databases and converted to VCF format. These were then annotated using above described pipeline and additionally automatically scored according to the ACMG 5-tier classification (Richards et al., 2015) using InterVar (Li & Wang, 2017). Variants scored as (likely) pathogenic were filtered. The allele count (AC) for each of these variants was normalized to the maximum allele number (AN) in the respective database and subpopulation. To estimate the carrier frequency for (likely) pathogenic *TECPR2* the normalized AC was added up and then divided by the maximum AN. Expected incidence was then calculated based on the estimated carrier frequencies and Hardy-Weinberg equation (see Supplementary File S2 sheets "gnomAD", "BRAVO" and "carriers" for details).

**Analysis of Missense Variant from Spanish water dog**

The literature search also identified a publication describing *TECPR2*-associated disease in Spanish water dogs (*Canis lupus familiaris*). (Hahn et al., 2015) The affected dogs showed gait abnormalities and behavioral deficits; together with histopathological results, these indicate a juvenile-onset neuroaxonal dystrophy. For comparability the therein identified *TECPR2* missense variant XM_005623828.3:c.4009C>T, p.(Arg1337Trp) was mapped from the dog reference genome "Broad CanFam3.1/canFam3" (chr8:g.70433803C>T) to the human reference genome GRCh37/hg19 (chr14:g.102964493C>T) using UCSC Browser and UCSC BLAT.

**Modelling of TECPR2 Protein Structure**

To analyze the missense variant distribution in the tertiary protein structure, we first searched the protein data bank RCSB PDB for crystal structures and ModBase for homology-based protein models. As this search identified no reliable model covering the identified *TECPR2* missense variants, we used the GalaxyWEB pipeline (Ko, Park, Heo, & Seok, 2012) to 1) detect protein modeling units using GalaxyDom, 2) predict the structure of these units from their protein sequence in FASTA format uploaded to GalaxyTBM (Ko, Park, & Seok, 2012) and 3) refine the top model structure using GalaxyRefine (Heo, Park, & Seok, 2013). In addition to the template-based modeling using GalaxyTBM/GalaxyRefine we also uploaded the determined modeling units to SWISS-MODEL (homology-modelling) (Waterhouse et al., 2018) and trRosetta (*de novo* protein structure prediction) (Yang et al., 2020). Resulting structures in protein data bank (PDB) format for all algorithms used in our analysis and for visualization (Figure 1) are provided in Supplementary File S3. Resulting structures were visualized with the Pymol molecular visualization software (Version 2.4.0b0; Schrödinger LLC,

New York, USA). We employed the mutation3D algorithm (Meyer et al., 2016) to search for spatial missense clustering in the generated structures.

**Analysis of Missense Variant Spectrum**

Distribution of the *TECPR2* missense variants was investigated visually by plotting them onto the secondary protein structure enriched with domain information from UniProt ("UniProt: a worldwide hub of protein knowledge," 2019) and comparing the distribution to missense variants reported as homozygous in gnomAD (Figure 1). We then analyzed protein regions constrained for missense variation by generating all possible *TECPR2* missense variants and by annotating them using the above described pipeline. Computational missense prediction scores were plotted by amino-acid position and a smoothed line was fitted using generalized additive models ("geom_smooth" function in ggplot2).

**RNA isolation**

The PAXgene Blood System (Becton Dickinson, Franklin Lakes, NJ) was used to extract RNA from peripheral blood lymphocytes of P1 and both parents. RNA was processed with DNase I (Qiagen, Hilden, Germany) and transcribed into cDNA with the Superscript II Reverse Transcriptase Kit (Invitrogen, Carlsbad, CA) according to manufacturer's protocol.

**RT-PCR analysis for monoallelic expression**

RT-PCR on cDNA from P1 and his parents was performed with two different primer pairs, each located in different exons and spanning the c.2829del, p.(Asn944Thrfs*7) *TECPR2* variant (P1f: 5′-ATCATCAGGACCAGTGGGGA-3′ and P1r: 5′-TGCCAGTTCTGAACCACAGG-3′ (amplicon length: 316bp); P2f: 5′-

AGTCACCATCAAGGGGAAGC-3′ and P2r: 5′ TCTCCTTGGGGCTTCTTGGA-3′ (amplicon length: 423bp)). Resulting PCR products were visualized by gel electrophoresis and subsequently Sanger-sequenced.

**RNA expression analysis by quantitative real-time PCR**

We analyzed RNA expression levels of *TECPR2* in P1, both his parents and two in-house control samples by quantitative real-time polymerase chain reaction (qPCR) with custom primers on an Eco48 qPCR cycler (NIPPON Genetics Europe, Düren, Germany). Quantification of two previously established qPCR primer pairs (*GAPDH*: 5′-CCACTCCTCCACCTTTGACG-3′ and 5′-CCCTGTTGCTGTAGCCAAATTC 3′ (amplicon length: 101 bp); *GUSB*: 5′-CAGAGCGAGTATGGAGCAGA-3′ and 5′-TATTCCCCAGCACTCTCGTC-3′ (amplicon length: 202 bp)) was to normalize the *TECPR2* expression (*TECPR2*: 5´-TGCTTGTGGGAAAGTCACCA-3´ and 5´-TCCCCACTGGTCCTGATGAT-3´ (amplicon length: 114 bp)) of each sample. Relative gene expression ratios were calculated using the Pfaffl method (Pfaffl, 2001) and normalized to those of the corresponding controls.

**Review of carrier tests**

A review of carrier screening test for individuals of Jewish descent was done via Google search with the terms “carrier screen tecpr2” on 2020-09-03.

### SUPPLEMENTARY FIGURES AND TABLES

#### Figure S1 | TECPR2 Structure with Additional Computational Scores for Missense Variants.

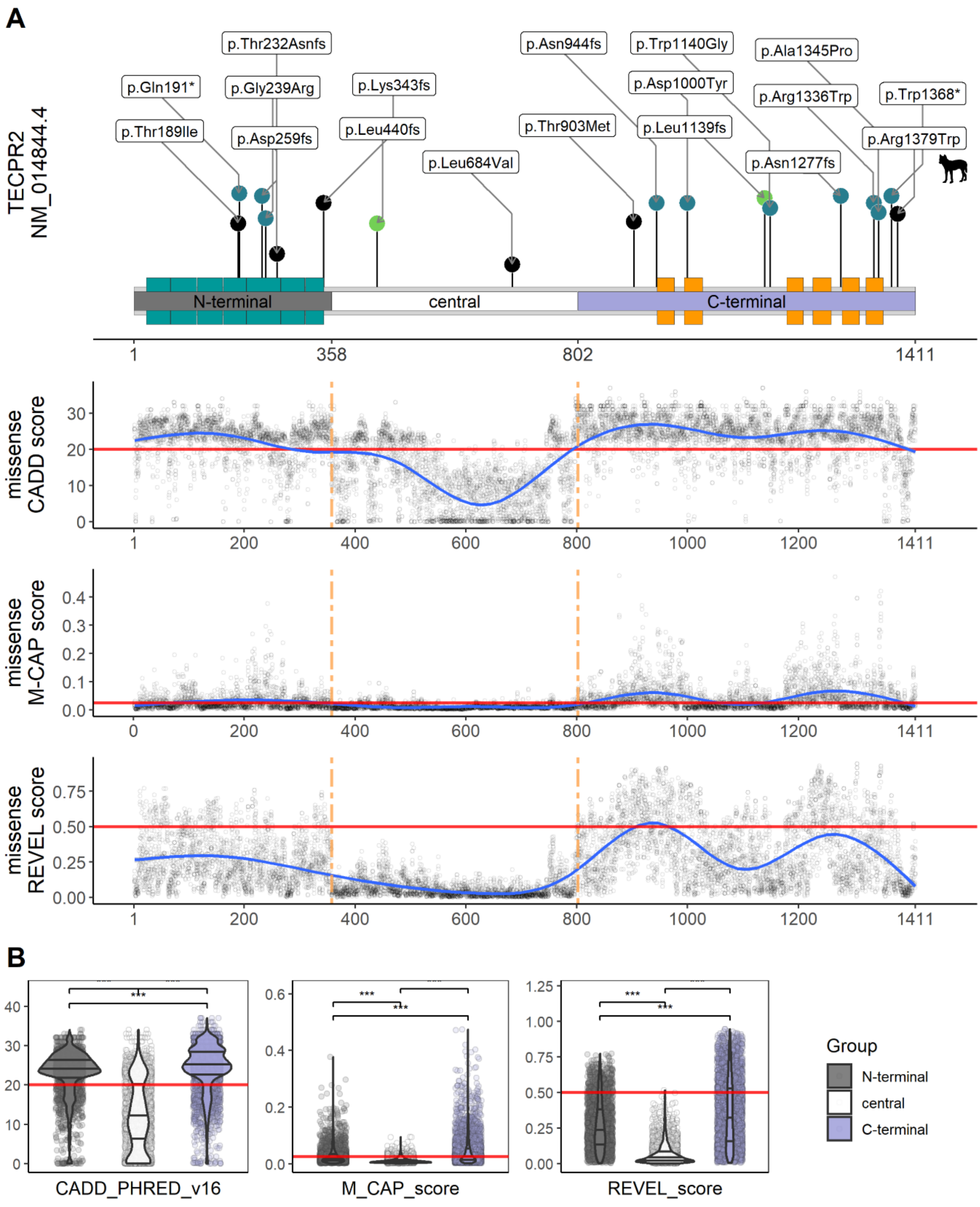


(A) Upper panel: Schematic of the TECPR2 secondary structure with WD40 and TECPR repeat units (WD40: green, TECPR: orange; based on Uniprot O15040) and three modeling units (“N-terminal”: grey, “central”: white, “C-terminal”:purple) identified by GalaxyDom and all variants depicted as lollipops as in Figure 2 with the variant from the Spanish water dogs added and labelled with a schematic dog. Generalized additive models comparing CADD PHRED v1.6 (second panel), M-CAP score (third panel) and and REVEL score (fourth panel), across the protein secondary structure for all possible missense variants in *TECPR2*. The red horizontal line marks the respective recommended cut-off for eachs core. The tan vertical dashed lines mark the borders of the modeling units. Note the srriking difference of missense score values between the three regions, which is especiall pronounsed for the CADD score. All three scores indicate 1) that the “central” region is least conserved, 2) higher conservation of the C-terminal region containing the TECPR repeats compared to the “N-terminal” region containing the WD40 repeats and 3) that the “C-terminal” region contains two conservation peaks (compare Figure S2). (B) Violin- and scatter-plot comparing the CADD PHRED v1.6 (left panel), M-CAP score (middle panel) and REVEL computational scores (right panel) for missense variants in the three modeling units. Missense variants in the three modeling units show significantly differenent scores for all three prediction algorithms recapitulating and confirming the visualization in A. Two-sided Wilcoxon signed-rank used for significant testing. ***: P<0.001. All data from File S3 sheets “TECPR2_domains“ and “allMissense”.

**Figure S2 | Comparison of Tertiary Protein Models generated.**

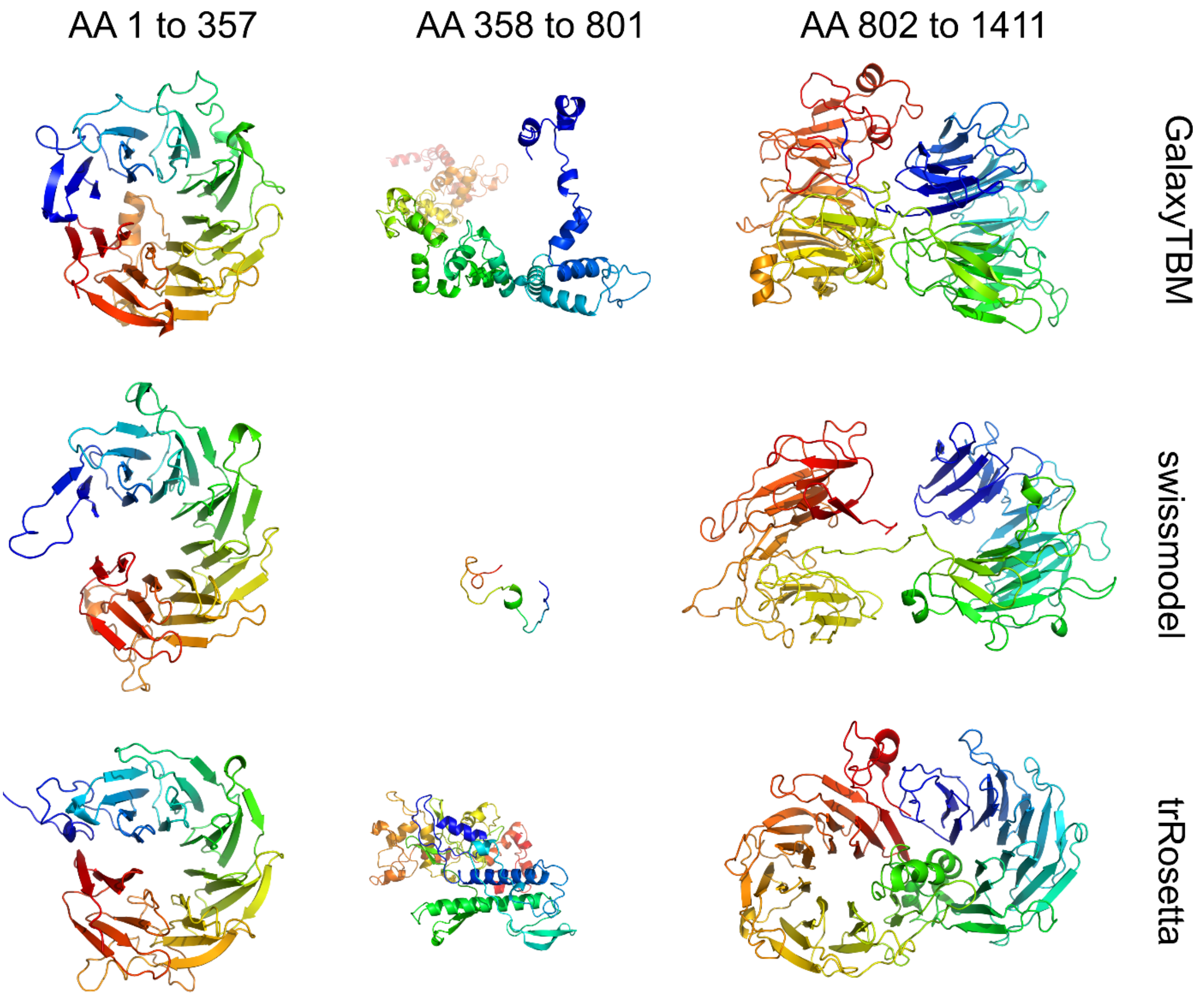


Representative PyMOL renderings (protein backbone presented as cartoon in rainbow colouring) of the top model for each of the three modelling units (columns: left: “N-terminal” AAs 1 to 357, center: “central” AAs 358 to 801, right: “C-terminal” AAs 802 to 1411) generated through the three algorithms/pipelines (rows: top: GalaxyTBM, middle: swissmodel, bottom: trRosetta). Compare also Figure 1B and C. Raw PDB files and protein FASTA files used as input for the algorythms are available as online dataset (Popp & Neuser, 2020).

Note the remarkably similar prediction of a circularised 7-bladed β-propeller structure for the “N-terminal” region containing the WD40 repeats. Also, the “C-terminal” region with the annotated TECPR repeats is predicted to form two β-propeller folds by all three algorithms; predictions mainly differ in the relative positioning of these β-propellers to each other. The central region instead shows extremely variable modelling results, with swissmodel only generating a 36 AA small model; this indicates an unstructured region in the middle of the TECPR2 protein which links the two β-propeller regions. Interestingly all missense variants with possible pathogenicity presented in our cohort or the literature affect AA residues in these conserved and structured regions. However clustering analysis of missense variants indicated no significant proximity of affected AA residues (Table S3), indicating that the AA substitutions do primary not hinder interaction but could rather cause structural changes in the protein which could lead to instability and faster degradation of misfolded proteins. Also, compare the overlapping results for the missense score analysis (Figure 1A; Figure S2). See Table S2 for groupwise root-mean-square deviation (RMSD) values for these models.

**Figure S3 | Results of qPCR for P1**

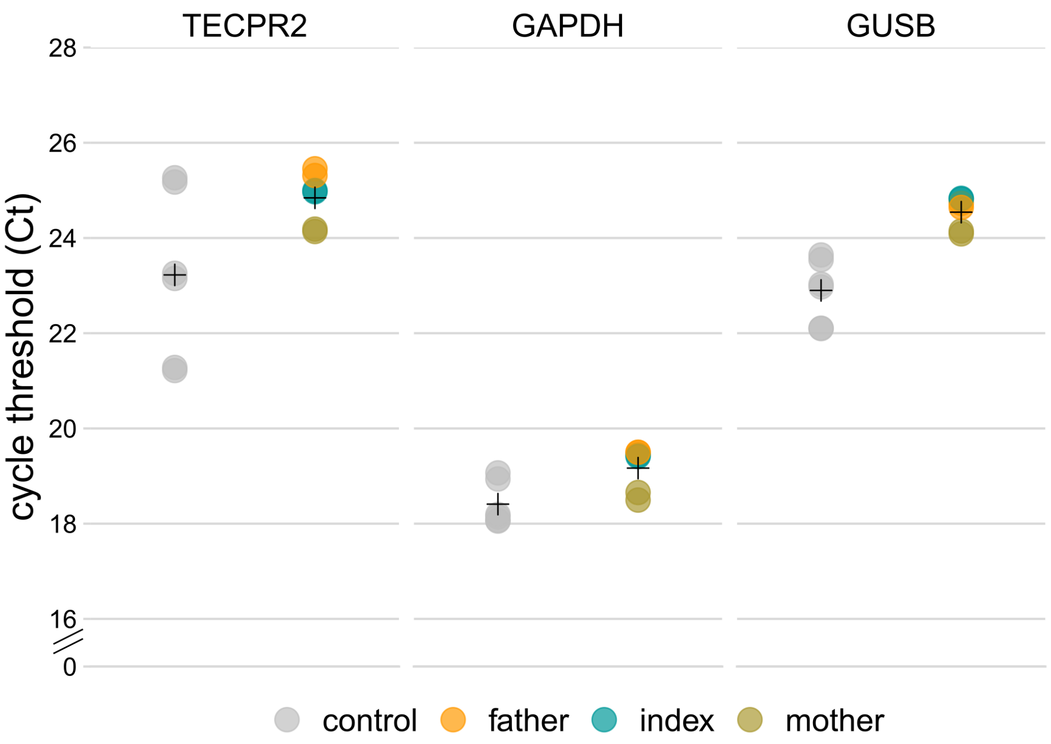


RNA expression levels of *TECPR2* and two in-house control samples (*GAPDH* and *GUSB*) in P1 (index) and both his parents (mother and father) compared to controls. Crosses show respective mean values of Ct.

**Figure S4 | Brain MRI and chest CT of P6**

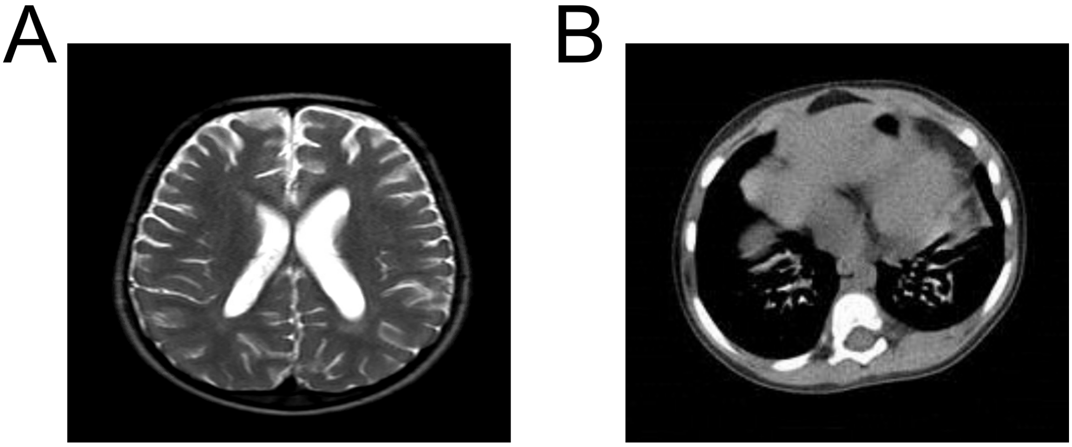


(A) T2 sagittal cranial MRI shows leukoencephalopathy with significant paucity of white matter with a spacious supratentorial ventricular system and prominent extra-axial CSF spaces. (B) CT chest shows multiple cystic and traction bronchiectasis changes in the basal segments of both lower lobes of the lung.

**Table S1 | Reviewed carrier tests**

| test | provider | website (accessed 2020-09-03) | TECPR2 included | listed carrier frequency |
|---|---|---|---|---|
| Expanded Carrier Screen (152 genes) | Sema4, Stamford, USA | https://sema4.com/products/test-catalog/expanded-carrier-screen-152/ | yes | 1 in 151 |
| Sephardi-Mizrahi Jewish Carrier Screen (54 Genes) | Sema4, Stamford, USA | https://sema4.com/products/test-catalog/sephardi-mizrahi-jewish-carrier-screen/ | yes | 1 in 151 |
| Ashkenazi Jewish Carrier Screen (64 Genes) | Sema4, Stamford, USA | https://sema4.com/products/test-catalog/ashkenazi-jewish-carrier-screen/ | no | 1 in 151 |
| Invitae Comprehensive Carrier Screen | Invitae, San Francisco, USA | https://www.invitae.com/en/physician/tests/60100/ | yes | NA |
| Inheritest Ashkenazi Jewish Panel | Integrated Genetics, Westborough, USA | https://integratedgenetics.com/inheritest-carrier-screen-ashkenazi-jewish-panel | no | NA |

**Table S2 | Root-Mean-Square Deviation (RMSD) Values for Protein Models.**

| **Model_1** | **Model_2** | **RMSD** | **Atoms** |
|---|---|---|---|
| GalaxyTBM_NP_055659_1_357_model_1_refine_model_1 | swissmodel_NP_055659_1_357_2020-07-28_model01 | 2.129 | 1920 |
| GalaxyTBM_NP_055659_1_357_model_1_refine_model_1 | trRosetta_NP_055659_1_357_TR014527_results_model1 | 3.588 | 2580 |
| swissmodel_NP_055659_1_357_2020-07-28_model01 | trRosetta_NP_055659_1_357_TR014527_results_model1 | 3.899 | 2107 |
| GalaxyTBM_NP_055659_358_801_model_1_refine_model_1 | swissmodel_NP_055659_358_801_2020-07-28_model01 | 9.998 | 273 |
| GalaxyTBM_NP_055659_358_801_model_1_refine_model_1 | trRosetta_NP_055659_358_801_TR014528_results_model1 | 51.663 | 3955 |
| swissmodel_NP_055659_358_801_2020-07-28_model01 | trRosetta_NP_055659_358_801_TR014528_results_model1 | 10.275 | 269 |
| GalaxyTBM_NP_055659_802_1411_model_1_refine_model_1 | swissmodel_NP_055659_802_1411_2020-07-28_model01 | 17.328 | 4318 |
| GalaxyTBM_NP_055659_358_801_model_1_refine_model_1 | trRosetta_NP_055659_358_801_TR014528_results_model1 | 17.846 | 5629 |
| swissmodel_NP_055659_802_1411_2020-07-28_model01 | trRosetta_NP_055659_358_801_TR014528_results_model1 | 12.538 | 3953 |

Pairwise RMSD values for all protein models (Figure S2 and (Popp & Neuser, 2020)) computed in PyMOL using the “extra_fit” alignment command.

**Table S3 | results of mutation3D clustering**

| Model | Input | Cluster_AAresidues | Cluster_Angstroms | Cluster_Pvalue |
|---|---|---|---|---|
| GalaxyTBM_NP_055659_1_357_model_1_refine_model_1.pdb | 189,239 30 357 10000 | NA | NA | NA |
| swissmodel_NP_055659_1_357_2020-07-28_model01 | 189,239 30 357 10000 | 189,239 | 14.7435 | 0.1373 |
| trRosetta_NP_055659_1_357_TR014527_results_model1 | 189,239 30 357 10000 | NA | NA | NA |
| GalaxyTBM_NP_055659_802_1411_model_1_refine_model_1 | 199,339, 544,578 30 610 10000 | 339,544,578 | 29.3262 | 0.2709 |
| swissmodel_NP_055659_802_1411_2020-07-28_model01 | 199,339, 544,578 30 610 10000 | NA | NA | NA |
| trRosetta_NP_055659_802_1411_TR014529_results_model1 | 199,339, 544,578 30 610 10000 | 199,339; 544,578 | 28.7559; 20.1757 | 1.956; 1.5524 |

Results of clustering analysis for missense variants in the conserved “N-terminal” and “C-terminal” regions using the mutation3D command line tool with amino acid residue positions of all reported possibly pathogenic missense variants mapped onto the predicted structure (Family E II-1: c.566C>T p.(Thr189Ile) → 189, In-03: c.715G>A, p.(Gly239Arg)) → 239; In-07: c.2998G>T p.(Asp1000Tyr) → 199, In-06:c.3418T>G p.(Trp1140Gly) → 339, In-03: c.4033G>C p.(Ala1345Pro) → 544, Dog: c.4135C>T (p.Arg1379Trp) →578). With 10,000 simulations and cluster size of up to 30 angstroms no significant cluster was identified in any of the models.
